# “If there is no motivation, the work stands still”: Lived experiences of local health managers innovating to retain health workers in the most deprived districts in Ghana

**DOI:** 10.64898/2026.02.04.26345577

**Authors:** Patricia Akweongo, Baatiema Leonard, Samuel Amon, Sedzro Kojo Mensah, Kassim Basit, William Akatoti, India Hotopf, Moses Aikins, Joanna Raven

## Abstract

**Background:** Ghana is facing health workforce crisis, with mass exodus of experienced health workers out of the country, and challenges in retaining them in deprived areas. The local initiatives of health managers and community leaders in successfully retaining staff in deprived areas are less documented.

**Objective:** Using Maslow five-tier motivation model (physiological needs, safety, love and belonging, self-esteem, self-actualization), we describe how local management initiatives motivate and support health workers to stay in deprived areas.

**Method:** This study employed an exploratory qualitative approach. In-depth interviews were conducted with 21 district health managers and nine focus group discussions with community leaders between February and March 2024 as part of a larger project aimed at improving health workforce retention in the three most deprived districts in the Eastern region of Ghana. Interviews were audio-recorded and transcribed verbatim. The thematic framework approach, supported by QSR Nvivo 17 was used to analyse the data.

**Results:** Local managers innovated using existing resources, relationships and networks to engage, motivate and retain health workers in deprived communities in line with Maslow 5 tiers of motivation. They observed that meeting physiological needs, such as having decent living conditions and adequate compensation to meet their daily necessities, was a prerequisite for motivating and retaining staff. Community leaders played a critical role in safety needs through providing accommodation and security to safeguard the facility. To meet love and belonging needs in such isolated areas, they provided a unique community welcome for newly posted staff, set up peer support platforms, provided family support, and enhanced school facilities for staff with children. Local managers addressed self-esteem by prioritizing and lobbying for remote health workers who were due for promotion and study leave. Self-actualization was less visible, but it was reflected in a manager stating, “If there is no motivation, the work stands still.”

**Conclusion:** Without deliberate efforts to motivate staff, it is challenging for managers to attract and retain health staff willing to work in and stay in deprived communities. Existing human resource policies to motivate staff should provide guidelines that focus on ensuring these five tiers are met for the retention of staff in deprived communities.

## Introduction

Rural and hard-to-reach areas present a unique challenge for retaining the health workforce. Globally, rural communities suffer disproportionately from chronic shortages of health professionals, as their needs are often unmet in these settings [1–3]. The World Health Organization (WHO) notes that half of the world’s population lives in rural areas but faces major difficulties accessing healthcare, largely because health worker density is far below that in urban regions [4,5]. Sub-Saharan Africa (SSA) has an estimated 2.5 health workers per 1,000 population, far below the WHO’s recommended SDG density threshold of 4.45 per 1,000 population [4]. A global shortfall of 10 million health workers by 2030 is projected, disproportionately impacting low-resource and rural settings and hindering health service delivery [6]. Achieving a transformed health workforce and addressing the availability and distribution of health workers in such settings is therefore key to improving health outcomes, especially in rural and underserved populations [3].

Ghana is facing a health workforce crisis, marked by the mass exodus of experienced health workers and by challenges in attracting and retaining health workers in underserved areas. Consequently, Ghana is unlikely to meet the Universal Health Coverage (UHC) target for key health indicators in 2030 [7,8]. In Ghana, at the end of 2018, there were 115,650 health sector staff, with 63.9% (74,250) of them being employed by the Ghana Health Service (GHS), 17% by the Christian Health Association of Ghana (CHAG), 10% by the teaching hospitals and 8.9% by other employers (5). The GHS annual report showed a health workforce density of 2.65 per 1000 population in 2017 [9]. In 2018, however, the WHO Regional Office for Africa estimated Ghana’s health workforce density was less than 2.22 per 1000 population (7). The inequitable distribution of the health workforce has also been identified, with fewer staff in rural, remote, and disadvantaged regions and communities. Achieving an equitable distribution of the Ghanaian health workforce remains a critical challenge, as studies and national policies [9]. The Government of Ghana has made efforts to address the inequitable distribution of the healthcare workforce in recent years. The 2020 National Human Resource Policy and Strategies for Health have specific objectives related to the attraction and retention of the health workforce and equitable distribution of health professionals serving in deprived or underserved locations/facilities [10]. At the health service delivery level, the GHS 2018 annual report indicates that despite several attempts to deploy health professionals, they refuse to accept postings to underserved areas (5).

Despite these efforts, deprived areas continue to face low retention - local, innovative initiatives are required. It is recognised that health workers tend to cluster in cities where professional opportunities, infrastructure and amenities (schools for children, spousal employment opportunities) are better [11]. Rural settings typically lack electricity, clean water, decent housing, and professional support, as per Maslow’s theory of motivation, where unmet physiological needs can deter workers from accepting or remaining in such posts [10]. In addition, these settings can be socially and professionally isolating, threatening the belonging and esteem needs of health workers who may feel cut off from peers, feel undervalued or get stuck without career advancement [12].

The context and specific needs of these areas can be conceptualized using Maslow’s theory of motivation, which outlines the five Tiers of human needs (self-actualization, esteem, love and belonging, safety and security, and physiological needs) [13]. According to this theory, lower-level needs must be partially met for higher-level motivators to have a significant influence on behavior change. Although this structure has been criticised as rigid, Maslow acknowledges that human needs are intersectional and an individual can pursue several needs simultaneously but argues that where deficits exist in meeting the lower-level needs, those deficits would dominate till they are addressed [14]. This aligns with the needs of staff working in deprived and hard-to-reach areas [13]. The current Ghana health workforce policies tend to focus on tiers one and two while health workers who are in deprived areas require that the five tiers be addressed holistically. Local managers, given their knowledge of context-specific challenges, are well-placed to address workforce issues, primarily where gaps exist in meeting the basic needs of health workers [15,16].

Although the literature and policies identify the workforce challenges in deprived, hard-to-reach areas, they do not describe what local managers do to retain healthcare staff in their districts [17,18]. This paper explores district health managers’ and community leaders’ initiatives to improve the retention of women and men health workers in three of the most deprived districts in the Eastern Region of Ghana – Afram Plains North and South and Akwapim North. These findings can support other managers navigating the challenges of retaining health workers in deprived areas.

## Methods

### Study design

This exploratory study employed in-depth interviews (IDIs) and focus group discussions (FGDs) to explore the perspectives and initiatives of health managers and community leaders on the retention of health workers in deprived areas. Documenting both health managers’ and community perspectives was crucial to understanding the views of both the community and the health workforce on facilitators and barriers to health workforce retention. The paper draws on baseline situational analysis data from mixed methods participatory action research study which co-designed, implemented and evaluated interventions to strengthen health workforce retention in deprived Ghanaian districts.

### Study settings

The study was conducted in three severely deprived districts of the Eastern Region of Ghana: Kwahu Afram Plains North (KAPN), Kwahu Afram Plains South (KAPS), and Kwahu East (KE). These districts are island communities, with extremely bad roads and communication networks and poor socio-economic indices. These districts were also selected because they are classified as the most challenging in terms of human resources and service delivery outcomes. Many posts are unfilled, with 36-50% of staff being posted to these districts but not taking them up. Many health workers are lower cadres, including community health workers, working in the Community-Based Health Planning and Services (CHPS) level. Doctor-to-patient ratios are very low; for example, in Kwahu Afram Plains North, there is one doctor per 70,000 people (according to communication with the district).

### Sampling and recruitment

A purposive sampling technique was adopted to recruit participants for FGDs and IDIs. In each district, we conducted three FGDs with CHMCs (Community Health Management Committee) members at the Community Health and Planning Services (CHPS) level: one functional CHMC, one non-functional CHMC, and one CHMC close to the district capital. Functional CHMCs are defined as CHMCS that actively engage with communities and the health service to give voice to the community and to mobilize resources to support and strengthen the implementation of CHPS and non-functional CHMCS are dormant in performing these roles [19]. In total, we conducted nine FGDs with CHMC members.

For IDIs, we purposively selected seven key stakeholders in each district, including health managers at the facility and district level, local government and community leaders (District Assembly members and chiefs). We conducted a total of 21 IDIs comprising three local government representatives, two traditional leaders, three District Directors of Health Services and 13 facility heads. These stakeholders were knowledgeable about the district’s health workforce situation. They provided valuable information on health workforce issues, retention policies, and initiatives to address workforce challenges. We considered gender in participant selection, even though, in such a deprived area, most managers and leaders were male (19 males and 2 females).

### Data collection

Data was collected between February and March 2024 using face-to-face interviews.

IDIs explored the perspectives of health managers and community leaders on factors that support or hinder health workforce motivation and retention in deprived areas, as well as the ways health workers are supported to stay in these districts. FGD topic guides covered the workforce situation in the district, reasons for shortage and retention of health workers, knowledge of attraction and retention policies, current strategies managers and community leadership use to address health workforce challenges and to support retention.

Interviews and FGDs were recorded once informed consent were obtained. In addition, the researchers took detailed notes. FGDs lasted approximately 1 hour and 15 minutes, and IDIs lasted 45 minutes, taking place in the health facility and community settings of the participants.

### Data management and analysis

The data (audio recordings) were transcribed verbatim. Unique codes were used to anonymize the data in such a way no one could trace the text back to the respondent. After anonymization, data from qualitative interviews and discussions were managed in NVivo 12 and analyzed using the thematic framework analysis approach (24).

We read the transcripts and developed a deductive coding framework based on the topic guides and research objectives, with new inductive themes iteratively incorporated into the coding framework. We then applied the framework to the transcripts and data and developed global categories and sub-themes based on the Maslow five tiers of motivation (self-actualisation, Esteem, Love and belonging, Safety and Security, and physiological needs). The research team then identified and agreed on key themes and illustrative quotations to support the tiers.

### Ethical considerations

Ethical clearance was obtained from the Ghana Health Service (GHS-ERC:007/02/24) and Liverpool School of Tropical Medicine (24-002). All participants were fully informed about the purpose of the study, the nature of their participation, and how the data would be used. Informed consent was obtained from each participant before collecting any data, ensuring that respondents knew participation was voluntary.

### Rigour and trustworthiness

To ensure the quality and rigour of the findings, several strategies were employed. Data collection tools were developed in English, translated into the predominant local language in the study districts, and then back translated into English to ensure accuracy and conceptual equivalence. During data collection, data collectors took detailed field notes alongside the audio recordings. At the end of each interview, data collectors briefly debriefed participants by summarising key points to confirm that the notes accurately reflected their views and to clarify any misunderstandings. Daily debriefing meetings were held between the data collectors and the research team to review emerging issues, refine probes, and ensure that the collected data were relevant, comprehensive, and consistent. All interviews were audio-recorded, and the recordings and detailed notes were reviewed promptly to identify any unclear issues that could be followed up and explored in greater depth in subsequent interviews or with other participants.

## Results

Maslow’s motivation framework is used to examine the multifaceted strategies needed to motivate and retain health workers in deprived districts by local managers. Where there is a lack of the necessities of life, physiological needs become a priority for staff. In Maslow’s hierarchy of needs, safety comes after physiological needs are met. The results show that in deprived areas where basic infrastructure and resources are lacking, the lower tiers of the motivation framework take prime importance. Local health managers intuitively recognized this and implemented a range of initiatives corresponding to each level of Maslow’s hierarchy to encourage health staff to stay (Figure 1).

**Figure 1.**
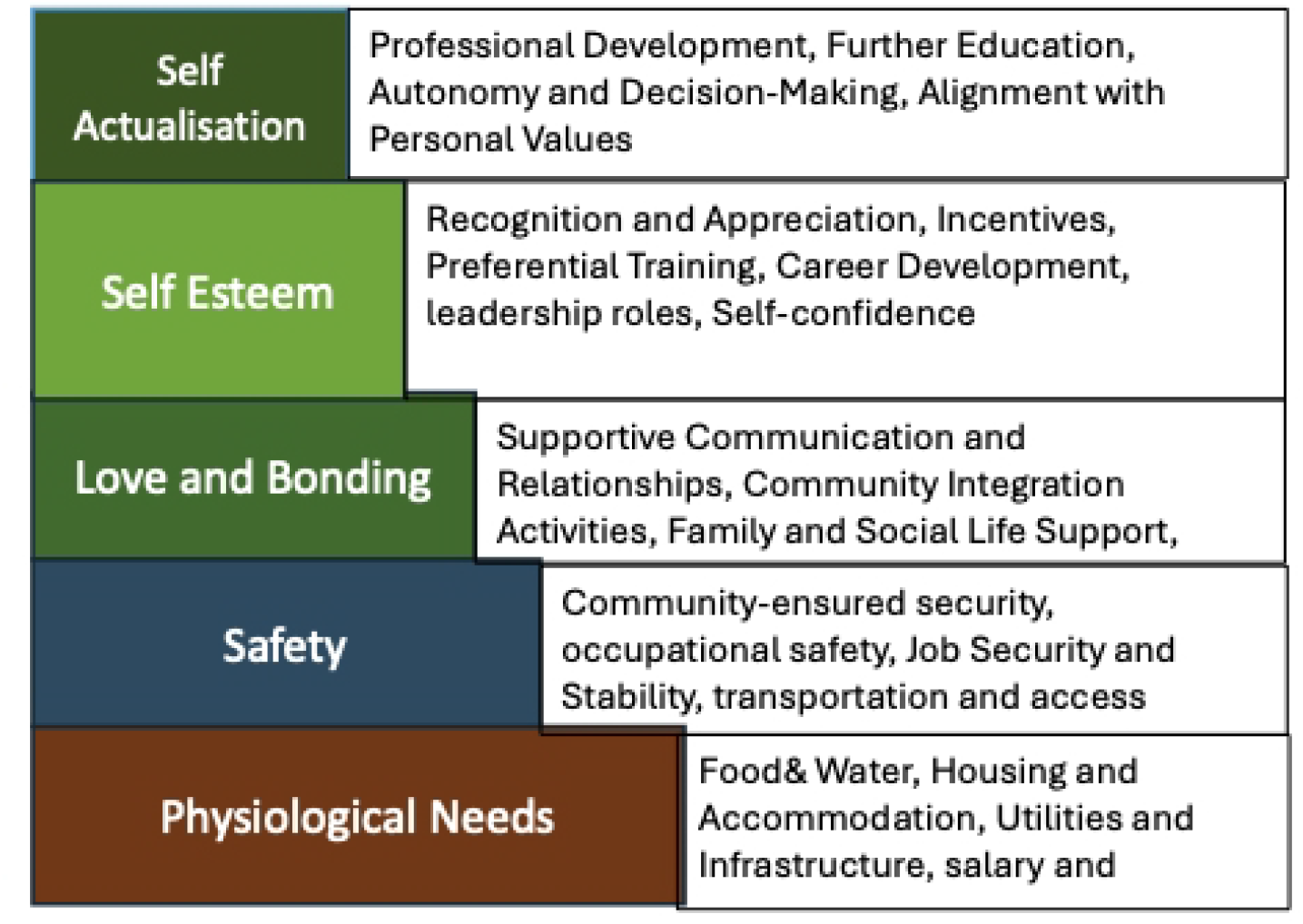
Overview of strategies to motivate health workers in deprived districts.

### Physiological Needs

The most basic physiological needs include food, water, shelter and other factors necessary for survival. For health workers, especially those accepting rural postings, physiological needs translated to having decent living conditions and adequate compensation to meet their daily necessities. In these deprived districts, local health managers observed that meeting physiological needs was a prerequisite for retention. They noted, for example, that newly posted health staff often faced long delays (six months to a year) before receiving their first salary, which in a remote setting with few financial reserves is a disincentive. To counteract this, managers went “out of their way” to support new staff with necessities until their pay came through, sometimes by expediting administrative paperwork through personal contacts in the bureaucracy so that employees could begin receiving income to cover food and living costs. Ensuring that staff do not go hungry or homeless was seen as the very first step.

Multiple initiatives were used by health managers to address the physiological needs of staff in deprived areas, and these were creative and community driven.

#### Provision of Food and Water

Local health managers and community members typically collaborated to ensure health staff had adequate food and water. In one district, the health facility acquired a plot of land to cultivate food crops and rear animals, generating foodstuffs that could supplement staff diets and even be sold to provide a small year-end monetary bonus to workers. Community volunteers in two districts also took it upon themselves to feed and provide for staff:

> “*I have taken it upon myself to feed them morning and evening, almost every day for about 18 months now. They have no problems in that regard*” *(KAPN, Community Volunteer, Female)*

Another volunteer explained that because the government does not pay new nurses for months after they are posted, villagers might contribute food from their farms to support them until their salaries arrive.

> *“Giving them foodstuffs tends to support them until they receive their salaries*… *they appreciate the support and warm welcome” (KAPS, Community Volunteer, Female)*
>
> *“Well, I advise my people to motivate them. Anything they get from the farm and they come, like foodstuffs and all those things, they can give it to them. And they have been doing it” (KE, CHMC leader, Male)*

Ensuring access to clean water was also a critical factor for staff. For instance, in KAPN, an NGO (World Vision) partnered with the community to install a water system featuring solar-powered pumps. In contrast, community members provided labor to ensure that the facility and staff housing had running water. In another case, a community leader recalled exempting health workers from paying for water at the communal standpipe and arranging for community members to deliver water to staff residences, making life a bit easier. These efforts directly supported the physiological needs of staff, thereby creating conditions under which health workers can work without experiencing food and water deprivation.

#### Housing and Accommodation

Housing and accommodation are basic needs that were often lacking or inadequate in rural postings. Communities and health managers stepped in to provide or improve accommodation for health staff as a strategy for retention. In some areas, community members built or upgraded health facilities with attached staff quarters.

> *“Through the effort of the community, they built a new CHPS center that is spacious and comfortable for the health staff to work in,”* said a volunteer *(KAPN, Community Leader, Male)*

Another manager explained that various communities took upon themselves to build houses so that incoming personnel have a place to live.

> *“Some have built one room, some two rooms, some have been able to build about four rooms*… *two for the facility and two for staff accommodation”. (KAPN, Manager, male)*

Renovation of existing buildings was also a common initiative among the community. For example, a community leader described renovating a “boys’ quarter” (outbuilding) to serve as nurses’ accommodation so that staff would not have to commute from far away. In one case, the chairperson of a CHMC even offered his own house to a newly assigned Physician Assistant so that they could live conveniently near the clinic. These anecdotes demonstrate a high level of local commitment to meeting the housing needs of health workers. From the managers’ perspective, having affordable or free housing available is a significant incentive for health staff. The communities helped satisfy a fundamental need, making it more acceptable for health workers to stay in deprived areas.

#### Utilities and Infrastructure

Healthcare practice in modern times relies on basic utilities such as electricity. Many of the hard-to-reach health facilities lacked grid power, which impeded patient care (e.g., the absence of refrigeration for vaccines and inadequate lighting for night duties) and also diminished the quality of life for staff (e.g., the lack of lighting, fans, and phone charging facilities). Local health managers, therefore, lobbied actively for electrification. In KAPN, the Health Director recounted making a “strong case” that every new facility should include provisions for water and electricity. They successfully attracted a partnership with GIZ (a German development agency) to install solar panels in remote clinics. Another manager from KAPS explained the impact:

> *“Here in the community, we did not have electricity so*… *some [staff] when they came, they left. But now, we have electricity and have also given them accommodation, as a result, when they come, they stay” (KAPS, Manager, Male)*

This implies *that* having electricity combined with housing is key to retaining staff. Noticing the lack of utilities in the facilities, communities became advocates, lobbying the government for the extension of the electrical grid to villages on isolated islands. Although challenging, such efforts demonstrate that community members recognized the power supply as key to retaining their health staff at the post. Electricity addresses both physiological need and also enables a safer, more functional working environment thus overlapping with safety needs as well.

### Safety and Security

For health workers in these three deprived districts living in isolated areas, safety needs centered around personal safety from crime and violence, occupational safety, secure housing infrastructure, job security and organizational support.

#### Community ensured Security

The communities in the three deprived districts played a proactive role in safeguarding their health workers. Community leaders and members organized informal security arrangements. For instance, in KAPN, where security threats were heightened, FGD participants highlighted that ever since a CHPS compound was established, *“there haven’t been issues of robbery or threats on [the staff’s] lives. They are safe here”*. While there were no official security guards, the community itself took responsibility for appointing community members to ensure the health workforce’s welfare and check on them.

Another community volunteer noted that, as their health facility was situated by the main road, they installed a lighting system around the clinic and recruited a community security person to watch over the facility at night. The assurance of security was cited as a key reason staff were comfortable staying, as explained by one community volunteer:

> *“ Because we have electricity, affordable accommodation and security is assured. That is why it is easy to stay in our community” (Afram Plains North, Community Volunteer, male)*.

Addressing the safety needs fosters a sense of trust. It reflects an understanding that retention is a two-way route, so that if workers risk serving in a deprived area, the community must also step up to make them feel secure and welcome.

#### Accessible Transportation

Safety and security were also viewed from the point of having accessible transportation. In these three deprived districts, difficult geography and poor roads posed safety hazards. Health staff had to travel long distances over rough terrain to reach their clinics or communities to provide healthcare services. Such journeys were not only exhausting but also risky, increasing the risk of breakdowns or robberies.

Local managers addressed this by prioritizing transport resources for remote workers. One district manager in Afram Plains North explained that when they receive new equipment like motorcycles from donors, they “*gave it to the staff on the island*” and took older motorbikes from the island staff to give to someone in a less remote area. The rationale was that those traveling through forests and rough paths *“needed good motorbikes even more. Their motorbikes keep breaking down*… *These are some of the little things I do to make them feel like I’m trying, though I can’t solve all their problems*.*”* (KAPN, District Manager, Male?)

This quote shows a manager’s empathy to reduce the risk and hardship of commutes for the most isolated workers. Additionally, communities engaged in maintaining roads and facility surroundings through communal labor, which was common across the 3 districts. Volunteers organized to weed and clear the clinic’s surroundings and access paths, making the environment more secure and presentable for staff and patients, as well as for service delivery.

#### Reliable Equipment and Supplies

Feeling secure in one’s job also means having the necessary tools to perform it without undue risk. In the three districts, many of the facilities did not have the basic equipment and apparatus to work with. Thus, where districts were able to provide a few new equipment, local managers prioritized the allocation of equipment and supplies to the remote posts. In IDIs with district managers, they mentioned replacing obsolete equipment for staff in deprived facilities, even if it meant those in better-endowed facilities had to use older ones. One manager captured it well, saying they deliberately

“discriminate” in favor of the hinterland staff to ensure they get more support, because “without deliberate effort to support staff in such remote areas, it is difficult… to get staff willing to accept work in such communities”. (KAPS,Manager,male)

### Love and Belonging

In Maslow’s hierarchy, love and belonging needs sit at the middle of the pyramid in which people yearn for friendship, family, intimacy, and a sense of connection after meeting physiological and safety needs. For health workers, especially those in hard-to-reach facilities far from their home, these needs are profound. Feelings of isolation, alienation, or lack of social support were frequently cited reasons for reluctance to work in remote areas or for leaving such posts.

To nurture a sense of belonging among their staff, particularly those in isolated outposts, one district director created a special WhatsApp group named “Director & Island” exclusively for the staff working on the distant island clinics. He explained, *“I have a general page for all staff, but I have a special group for my island people*… *I even call them when I don’t call other staff, just to check on them so they feel like I’m there for them, “*His colleagues teased him, calling him “the island director” because of how attentive he was to that subgroup, but he felt it was necessary *“to convince them [to stay], even to the extent that I have to go there with them to the facility*.”(KAPN Manager, Male) This hands-on approach by leadership significantly boosts belongingness.

#### Family and Social Life Support

Many health workers were hesitant to go to rural and deprived areas due to concerns for their spouses finding work and their children’s schooling, with many female health workers having to live far from their partners. District managers recognized this and implemented specific measures. For instance, in KAPS, it was noted that “staff who have children of school-going age tend to leave the community” unless there are decent schools. In response, the local health management partnered with the community to improve the local school by renovating school buildings and lobbying for more teachers so that health workers’ children could get a good education right there in the community.

### Esteem

Esteem refers to an individual’s desire for respect, recognition, achievement, and self-worth. There are two types of esteem needs with one being the need for external recognition (respect and appreciation from others, status, prestige) and the other being the need for self-esteem (confidence, personal sense of accomplishment). In the health workforce context, esteem can be translated to being recognized for good performance, having opportunities to advance or take on greater responsibilities, receiving titles or promotions, and feeling that one’s work is essential and valued. In the three districts, local managers made conscious efforts to boost health workers’ esteem, to encourage them to remain in deprived areas, using several initiatives.

#### Recognition and Appreciation

Local health managers understood that a simple pat on the back or a formal commendation can go a long way in making staff feel valued. Managers lobbied NGOs to provide special rewards, such as monetary incentives to doctors who stayed for one year in deprived districts. Community leaders also often express gratitude to health workers, publicly acknowledging their service during community meetings or events, which boosts the health workers’ self-esteem. One district health director explicitly stated that intangible motivators matter, emphasizing that little opportunities or gestures can make remote staff “feel like it’s something” …..”they may not be big but the little things you do… they feel like it’s anything. This came as a district manager in the Afram Plains North described favoring island staff for training slots, implying that those opportunities served as recognition for them.

#### Incentives Tied to Service

Some districts devised tangible reward schemes that confer esteem. For example, in KAPN and Kwahu East, they formed a partnership with an NGO called “Together for Ghana” and a philanthropist to provide a significant monetary bonus to any doctor who completed one full year of service in the district. This incentive addressed financial needs, but it was framed as a form of recognition for perseverance and may also build self-esteem as the doctors prove to themselves that they met the challenge.

#### Preferential Training and Career Development

To boost their esteem, district managers gave some health workers opportunities for growth and advancement. Local managers were very intentional about this for their rural staff. They prioritized remote workers for training programs and workshops as much as possible. One manager from the revealed that *“If for a training*… *you were requesting perhaps five people; I would have selected more of my island people than inland. That is what I do, so that they get the opportunities*… *For the programs that we do at the district level, I give those in the hard-to-reach areas more transportation [allowance] than those in the inlands. Of course, they deserve it*.*”* This demonstrates attempts to fulfill self-esteem needs as by giving these extra opportunities it built their skills, and it allowed them to feel noticed and acknowledged. Similarly in the Afram Plains South, health providers were offered more opportunities to participate in management training to enhance their skills as sub-district heads.

#### Self-Confidence

In the Afram Plain South and Kwahu East, managers helped build self-confidence by making sure those in remote areas got refresher trainings and were not left behind technically. The special attention and resources gave them confidence that they could manage even though they were working in remote areas.

### Self-Actualization

At the height of Maslow’s hierarchy is self-actualization, where the individual achieves one’s full potential, to grow, and to find fulfilment and meaning in what one does. For a health worker, self-actualization might translate into the desire to achieve one’s career goals, to innovate and solve challenges, to assume autonomy and leadership, to continue learning at the highest levels, or to contribute significantly to society. In the three deprived districts self-actualization featured less as local initiatives by managers focused first on getting the basics required to retain staff. However, it was observed that health workers had expanded roles and the autonomy to lead and make decisions.

#### Meaningful Work and Expanded Roles

A core aspect of self-actualization is feeling that one’s work is meaningful and utilizes one’s talents. District managers helped enhance this by ensuring health workers in remote posts had the needed tools to actually practice effectively (e.g., equipment, apparatus). They also, allowed these workers a broad scope of practice. For instance, a midwife learned to manage common illnesses or run a small laboratory test, practicing beyond a narrowly defined role. This can be enriching for such healthcare staff professionally.

Autonomy and Decision-Making: District health managers involved health staff and gave them lead roles to manage local initiatives. This helped staff to realize their leadership potential earlier in their careers.

However, intrinsic motivation likely played a role for those who stayed, as some health workers found personal fulfillment in the challenge and purpose of rural service. As one manager’s quote encapsulated, “If there is no motivation, the work stands still,” and some of that motivation for individuals can come from an internal drive to serve, leading to self-actualization.

## Discussion

This study examines how Maslow’s hierarchy of needs can inform strategies to motivate and retain healthcare workers in underserved districts in Ghana. Local managers employed several initiatives, recognizing that motivation is inherently multidimensional. Local managers, in collaboration with communities, implemented a range of initiatives like securing food, water, and decent shelter for staff (Physiological needs); improving security, transportation, and working conditions (Safety needs); fostering a welcoming and supportive community (Belonging needs); providing recognition, extra training, and incentives that made staff feel valued (Esteem needs); and finally, they enabled professional growth and meaningful work, laying the groundwork for personal fulfillment in service (Self-Actualization needs). These efforts, though born out of practical necessity, align closely with global best practices and underscore a universal fact that when health workers feel cared for, they can better care for others[20].

Improving the working and living conditions of health workers has been highlighted in several studies as necessary in retaining staff in deprived and hard-to-reach areas [21]. The three districts addressed this issue by installing solar panels, water systems, and new clinics and quarters, similar to how other countries have invested in rural infrastructure. For instance, Brazil’s successful Programa Mais Médicos (More Doctors Program) not only placed doctors in rural areas but also invested in upgrading primary care facilities and housing to support them (28). In many African countries, lack of basic infrastructure is the top complaint by rural staff *[21]*. Evidence from a discrete choice experiment in Ethiopia, for instance, showed health workers would trade off some salary for better facility amenities and career development prospects [22].

In low-resource settings, community and local leadership initiative is key to motivating and retaining health workers. This study reveals that empowering local managers and communities to innovate can yield context-specific solutions (like farming for food or lobbying an NGO for solar panels). It further suggests that mobilizing community support (e.g., providing housing, security) can significantly augment formal strategies. Similar findings are reported in Nepal, where local villagers have built homes for midwives, or in parts of India where Panchayats (village councils) provide stipends or honour ceremonies for doctors [23]. However, these are often ad-hoc. In Ghana and other African contexts, the health system leverages community support as community structures are often strong in rural Africa [24–26]. In contrast, a study found that a rural Chinese township hospital is fully government-run and the community has less role in providing housing or food. Thus, who provides support would differ depending on the context. In Ghana and across most of Africa, community and local government can collaborate to support health workers. In contrast, in many Asian countries, the opposite is observed: the central or provincial government provides standardized support [27]. This bottom-up innovation in supporting health workers is less documented globally, as it is somewhat unique in explicitly researching local retention initiatives to support health worker retention, as this study attests. The documentation of community contributions provides evidence to policymakers that community engagement could be systematically considered as part of retention policy.

The findings of this study also highlight retention challenges common across Sub-Saharan Africa, including low health worker densities, infrastructure deficits, and resource constraints. Similarly, the strategies in these countries often rely on donor funding or community efforts because government budgets are limited. For example, in Malawi and Zambia[28,29]. Large salary top-ups for rural workers were funded by donor programs in the 2000s, whereas in this study, in the three deprived districts in Ghana, we saw NGOs contributing in-kind to support physiological and safety interventions, which are critical needs in deprived areas in Ghana and other African settings. In many other places, where these physiological and safety basics are a given, the challenge lies more in intangible factors, such as professional isolation or a lack of educational opportunities. Thus, even in contexts where the tangibles are fixed, the intangibles remain critical issues requiring intervention. For instance, certain remote indigenous communities in Canada or Australia face issues of social isolation and cultural differences similar to those in Africa (health staff feeling isolated, etc.) [30,31]. Similarly, Saudi Arabia has modern clinics in rural areas but struggles to keep doctors there because they feel isolated and disconnected from city life, so they have to give extra pay and also arrange rotations or other measures [32]. So, the belonging need is universal, irrespective of the context and cultural differences even though the physiological and safety gaps are still very large in Ghana and Africa, requiring inputs of infrastructure and basic goods.

The study further demonstrates that no single intervention is key to addressing retention issues, necessitating a combination of context-specific strategies. [33,34]. The approach of local managers in this study offers a bundle of interventions that address multiple needs simultaneously, similar to global recommendations. For example, a Cochrane Review on interventions for increasing health workforce found that multifaceted interventions were more likely to show effect but taking into account the context [35]. The local managers in this study took a holistic approach, touching all Maslow tiers, showing why the initiatives had some success. Evidence shows that some other countries have tried one dominant strategy at a time. For example, financial incentive-heavy approaches like that of Papua New Guinea’s large rural allowance or Zambia’s rural hardship bonus had partial success but did not solve the problem on their own [36,37]. Conversely, in India, education/regulation approaches to support bonding without good incentives led to begrudging compliance among staff, followed by resignations [38].

The study also demonstrates that health workers who remain in deprived areas have intrinsic motivation to serve. For example, in this study we found a subset of health workers who remain in rural posts due to personal values or a sense of duty. These individuals often cite patient need, community attachment, or professional mission as reasons to stay, even when conditions are tough. For example, some nurses feel a commitment to serve similar communities, deriving personal fulfillment (self-actualization) from it. This is similar with the concept of “mission-oriented” health workers such as doctors serving in remote parts of Alaska or rural missionaries in Africa who are driven by altruism [39]. However, these health workers may still need to receive non-financial recognition so that those who are motivated by service can still feel valued by the system.

## Limitations of the study

This study sought the views of local health managers who included district health directors and sub-district heads and community leaders. Frontline health workers were not included as this was considered managerial, though their views could have provided additional information on their satisfaction with the initiatives. Despite seeking maximum variation for gender, we were unable to achieve a gender balance, potentially overlooking gendered dimensions of health workforce retention.

## Conclusion

Motivating and retaining health workers in rural and difficult-to-reach areas is a complex challenge, but as this study has shown, Maslow’s five-tier theory of human motivation offers a powerful lens to understand and address the holistic needs of the health workforce. By viewing health workers with a hierarchy of needs from the most basic physiological requirements to the deepest aspirations of self-actualization, health systems can design more effective, humane, and sustainable strategies to attract and keep staff where they are most needed.

## Author’s contributions

The Health Workforce retention was designed by JR and PA in collaboration with MA, LB and SA. The manuscript was initiated and written by PA, and reviewed by JR, LB and IH. Field work for this initiative was organized by PA, LB, SA, SM, ABK, WT, MA, JR and IH. Initial data analysis was conducted by PA, JR, IH, LB, SA, ABK, WA, SM and MA.

## Acknowledgements

We wish to acknowledge the Eastern Regional Health Directorate, the participating districts, and the Country Advisory Group Members for providing direction on the areas to focus on in this project. We also thank all our study participants.

## Data availability statement

The dataset is available from the lead author on request.

